# Working from a distance: Who can afford to stay home during COVID-19? Evidence from mobile device data

**DOI:** 10.1101/2020.07.20.20153577

**Authors:** Christine Dimke, Marissa Lee, Jude Bayham

## Abstract

As the COVID-19 pandemic continues, local and state governments must weigh the costs and benefits of social distancing policy. However, the effectiveness of such policies depend on individuals’ willingness and ability to comply. We propose a simple method to infer sociodemographic heterogeneity in social distancing as measured by Safegraph mobile device data. We document evidence that people’s ability to work from home is a determinant of time spent at home since the beginning of the pandemic. On April 15th, census block groups that are more likely able to work from home spent 3 more hours at home compared to those who were not. We see supporting trends among block groups with differences in income and educational attainment.

**JEL:** J19, J69, Z00

## 1. Introduction

The extent to which people reduce potentially infectious contacts is a function of economic conditions because people employed in essential positions may not have the flexibility to work from home. Baker et al. (2020) estimates that 18.4% of the US population works in occupations where they are exposed to COVID-19 at least once per month. These jobs tend to pay lower wages and are disproportionately held by minority populations, or people with lower educational attainment (Mongey & Weinberg, 2020). Additionally, households at lower percentiles of earnings experience larger drops in income during recessions (Heathcote et al., 2009). The dependence on this income makes it difficult for individuals to choose to stay home.

Occupation is a key determinant of who can stay home during the pandemic response. Dingel & Neiman (2020) identify which occupations have high and low ability to work from home. Workers in high-personal-proximity occupations with low-work-from home ability are less likely to have college degrees and less likely to be white. They are more likely to have below median income and more likely to work in small firms (Mongey & Weinberg, 2020). These small firms are less likely to remain open after crises (Mongey & Weinberg, 2020).

While useful, this static analysis does not indicate who has and who has not been distancing during this pandemic. Here we propose a simple method for parsing anonymized mobile device location data by sociodemographic characteristics using publicly available Census data. Our method yields up-to-date estimates of time spent at home across demographic groups, a classification unavailable using mobile device data alone. Our analysis extends the work of Jay et al. (2020), who document heterogeneous mobility by income quintiles, by evaluating education levels and occupations with the ability to work from home.

## 2. Materials and Methods

Our objective is to estimate social distancing by socioeconomic and demographic characteristics. We merge Census Block Group (CBG) level mobile device data from SafeGraph along with demographic data from the US Census Bureau’s American Community Survey (ACS) (U.S. Census Bureau, 2016). Safe-Graph (www.safegraph.com) aggregates anonymized mobile device data that can be used to understand movement patterns during the COVID-19 epidemic. To enhance privacy, SafeGraph excludes census block group information if fewer than five devices are observed on any day.

The CBG level data is granular, but also preserves device anonymity. There are over 200,000 census block groups in the US with an average population of just over 1500 (U.S. Census Bureau, 2016). We classify each CBG based on the composition of the population along the following characteristics: education, household income, and occupations with ability to work from home. Specifically, we identify CBGs with a majority of the population in one category. The education classification is based on two levels of education, those who have a Bachelor’s degree or higher and those who do not. The household income classification is based on three income brackets: $0-50,000, $50,000-100,000, and greater than $100,000. We assigned groups to having high ability to work from home or low ability based on the classification in (Dingel & Neiman, 2020). Specifically, we multiply the fraction of workers in each occupation that are likely able to work from home (Dingel & Neiman, 2020) by the number of people employed in each occupation, and sum this product across all individuals in the CBG.

We classify each census block group as dominated by a category of interest if the fraction of the population in one of those categories exceeds 51%. Not all census block groups have a dominant population, so these estimates are based on a subset of the sample. We run the analysis for thresholds between 50% and 80%. While the ordinal rank of time spent at home remains constant, the sample size falls as the thresholds are increased resulting in larger standard error estimates. Non-binary categories remain qualitatively stable between 40% and 70%.

We implement our social distance parsing approach via regression of the following form,

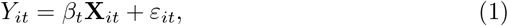

where *Y*_*it*_ is the vector of median time spent at home in CBG *i* on day, *t*, and *X*_*it*_ is the matrix of categorical dummies in a given regression. This model does not include an intercept, so *β*_*t*_ is interpretable as the group mean on day *t*. In contrast to the pre-post approach used in Bushman et al. (2020), this analysis allows us to see trends based on actual CBG data instead of hypothetical differences. We estimate *β*_*t*_ for each day from January 1 to June 23, 2020.

We can apply this methodology to any demographic variable available from the ACS. We omit three demographic variables from our report to succinctly describe results related to who can work from home. These variables are gender, age, and race, and they influence our understanding despite their omission here. These omitted categories are correlated with education, income, and employment status. We choose to keep this analysis separate rather than including controls because of barriers to identification and a recognition that such an analysis is biased.

## 3. Results and Discussion

Table 1 describes the variables constructed for the analysis. Column 1 displays the number of CBGs classified in each group while column 2 displays national mean values for each category. If people sorted into CBGs based on these characteristics, column 1 and 2 would match. We find that the fraction of CBGs dominated by households under $50,000 is comparable to the fraction of households. We find the no college degree and cannot work from home categories to be overly represented relative to national distributions. These results suggest that groups in lower socioeconomic categories are more clustered than other groups.

**Table 1:** Summary statistics of ACS and SafeGraph data.

| Category | CBG Count | CBG Means |
| --- | --- | --- |
| **Income (1000s)** | (%) | Households (%) |
| Under 50 | 92,860 (43.5) | 247.5 (47.8) |
| 50-100 | 6,952 (3.26) | 160.8 (29.3) |
| Above 100 | 22,410 (10.5) | 131.5 (23.0) |
| Not Classified | 91,274 (42.8) | NA |
| **Education** |  | Adults over 25 (%) |
| No College degree | 177,251 (83.0) | 683.9 (71.4) |
| Bachelors or Higher | 32,970 (15.4) | 296.6 (28.6) |
| Not Classified | 3,275 (1.53) | NA |
| **Work From Home** |  | Working Adults (%) |
| Cannot work from home | 186,774 (87.5) | 440.2 (66.8) |
| Can work from home | 21,182 (9.92) | 236.3 (33.2) |
| Not Classified | 5,540 (2.60) | NA |

We plot regression coefficients representing average median time spent at home by sociodemographic category. In all cases, we find that time at home declines throughout the first two months of the year. Importantly, there is little difference between categories prior to the pandemic. While the first case of COVID-19 was reported in late January, large-scale response began in late February and early March. Time at home increased throughout March and into mid April, after which states began reopening and time at home began to fall toward pre-epidemic levels. Around mid-May, the trend of time at home stabilized around mid-March levels. Notably, the difference between categories within education, income, and ability to work from home have declined since mid-May.

We explore the heterogeneity of this response along education, income, and ability to work from home (Figure 1). We find that those with Bachelor’s degrees or higher, household incomes greater than $100,000, and a greater ability to work from home spent significantly more time at home relative to the rest of the population. On April 15th, the initial peak of the COVID-19 response, people in occupations with higher ability to work from home spent approximately 3 more hours at home than those in occupations without the ability to work from home. As states and municipalities have reopened, people in all sociodemographic groups are spending less time at home.

**Figure 1:**
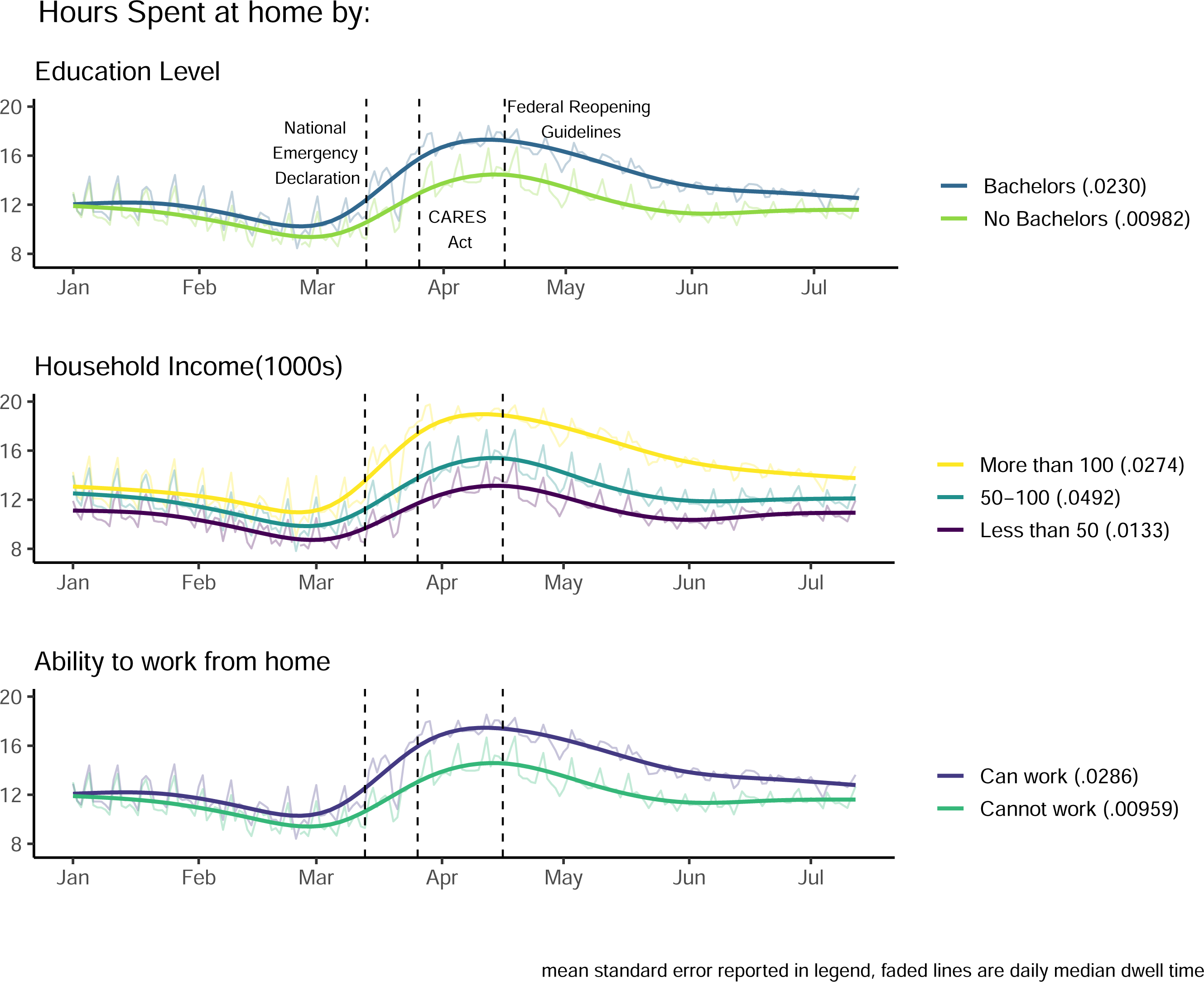
Time at home by a) educational attainment, b) household income, and c) ability to work from home. Vertical lines indicate: March 13th, the date President Trump declared a national emergency, March 27th, the signing of the Coronavirus Aid, Relief, and Economic Security (CARES) Act, and April 16th, the day the White House released plans for reopening the economy. The mean of the daily standard error estimates are reported in parentheses in the legend.

This simple approach to parsing distancing metrics has its limitations. It is possible that the population of device users in a CBG does not align with the category that it has been assigned in our analysis. Mobile device ownership is unequally distributed across society with younger people more likely to have smartphones and access to the internet. In 2018, 95% of individuals ages 18-34 in the U.S. had smartphones while only 67% of individuals older than 50 owned smartphones (Silver, 2019). Our classification method may be more robust to these effects since we focus on more homogeneous CBGs. However, more research is needed to understand the extent to which mobile device users are representative of the population at large.

## 4. Conclusion

Our analysis has several implications for social policy during the COVID-19 response. Those who are able to work from home spend significantly more time at home relative to their less able counterparts. If this at home work is as productive as the work that they performed under pre-COVID-19 circumstances, there is little to be gained by this portion of the population returning to work as usual. Employers can complement public health policy by working hard to accommodate at risk populations.

## Data Availability

As stated in the manuscript Safegraph compilation of ACS data is available for download free of charge. The mobility data can be licensed from Safegraph. Data processing code can be found in the repository linked.

https://github.com/cdimke/work_from_home

## 5. Acknowledgements

We acknowledge support from Amazon Web Services Diagnostic Development Initiative. We thank SafeGraph (www.safegraph.com) for generously providing data.

## Notes

Declarations of interest: none

### Competing Interest Statement

The authors have declared no competing interest.

### Funding Statement

This research did not receive any specific grant from funding agencies in the public, commercial, or not-for-profit sectors. We acknowledge support from Amazon Web Services Diagnostic Development Initiative. We thank SafeGraph (www.safegraph.com) for generously providing data.

### Author Declarations

No approvals were obtained due to the secondary nature of the data they were unnecessary.

